# Characterizing Acute Low Back Pain in a Community-Based Cohort

**DOI:** 10.1101/2023.10.02.23296149

**Authors:** Colleen Burke, Kenneth A. Taylor, Rebecca Fillipo, Steven Z. George, Flavia P. Kapos, Stephanie Danyluk, Carla A. Kingsbury, Kelley Seebeck, Christopher E. Lewis, Emily Ford, Cecilia Plez, Andrzej S. Kosinski, Michael C. Brown, Adam P. Goode

## Abstract

Acute low back pain (LBP) is a common experience, however, the associated pain severity, pain frequency, and characteristics of individuals with acute LBP in community settings have yet to be well understood. In this manuscript, three acute LBP severity categorization definitions were used based on LBP frequency combined with either 1) pain impact frequency (impact-based) or 2) pain intensity (intensity-based), as well as LBP pain interference frequency (interference only-based) severity categories. The purpose of this manuscript is to describe and then compare these acute LBP severity groups in the following characteristics: 1) sociodemographic, 2) general and physical health, and 3) psychological. This cross-sectional study used baseline data from 131 community-based participants with acute LBP (<4 weeks duration before screening and ≥30 pain-free days before acute LBP onset). Descriptive associations were calculated as prevalence ratios for categorical variables and Hedges’ *g* for continuous variables. Our analyses identified several large associations for impact-based and intensity-based categories with global mental health, global physical health, STarT Back Screening Tool risk category, and general health. Larger associations were found with social constructs (racially and ethnically minoritized, performance of social roles, and isolation) when using the intensity-based versus impact-based categorization. The interference-based category did not capture as much variability between acute LBP severity categories. This study adds to the literature by providing standard ways to characterize community-based individuals experiencing acute LBP. The robust differences observed between these categorization approaches suggest that how we define acute LBP severity is consequential; these different approaches may be used to improve the early identification of factors potentially contributing to the development of chronic LBP.

## Introduction

Up to 25% of individuals experience acute low back pain (LBP) annually.[48; 52] Despite being relatively common, acute LBP receives little attention due to perceived favorable long-term outcome relative to chronic LBP.[38; 45] However, the transition from acute to chronic LBP may be higher than previously thought, with 32% in a large cohort of US adults seeking care for acute LBP.[51] Additionally, it is typical for studies estimating chronic LBP incidence to use cohorts restricted to patients seeking healthcare, yet a large proportion (42%) of individuals experiencing LBP do not seek care.[21] Because of this, results from care-seeking cohorts may not be generalizable to adults with acute LBP.

Cohorts recruited directly from the community (i.e., not limited to individuals seeking care) play an essential role in understanding the full spectrum of health and disease processes regardless of healthcare access or utilization. For example, a recent Australian community-based cohort study indicated that acute LBP’s prognosis is far better than in clinical populations.[15] However, the same may not be accurate in the US, given the differences between these countries (e.g., demographic makeup, sociopolitical, and healthcare characteristics). Like studies of those seeking care for LBP, the limited US community-based studies available also indicate that chronic LBP incidence is higher than previously thought (approximately 25%).[25; 52]

More studies enrolling participants directly from the community are needed to understand acute LBP, and ultimately how these characteristics of acute LBP contribute to chronic LBP by means of transitioning from acute to chronic LBP. Recently developed measures to characterize chronic LBP focus on frequency-based questions to better understand high-impact chronic pain.[14] Prior studies discussing pain distributions have focused exclusively on categorizing the severity of chronic pain and examining the natural history of chronic pain.[18; 54] Various cutoffs for pain intensity (e.g., >4/10 on an 11-point scale) have been proposed to determine eligibility for LBP studies with the rationale that excluding individuals with lower pain intensity may provide greater scope for meaningful change in pain scores.[35] The downside is that studies applying such restrictions may miss important information regarding characteristics of the overall group of acute LBP sufferers such as key sociodemographic, general health/clinical characteristics, health behaviors, social health, and psychological factors.[34; 35] Recently, Eccleston et al proposed a framework for the examination of acute and chronic pain as well as the transition between states of pain.[19] However, to our knowledge, the empirical description of acute LBP subgroups of severity using concepts from this recently proposed framework has not been reported in the literature. To fill these gaps, we proposed three definitions to categorize the acute LBP experience that align with the recently proposed framework[19], in a US-based cohort study of participants recruited from the community. By taking into account acute LBP subgroup characteristics and quantifying effect sizes, we may provide further support for such a framework and improve the prediction of chronic LBP or the identification of factors that contribute to its development. The purpose of this manuscript is to categorize the severity of acute LBP and characterize these three subgroups by: 1) sociodemographic factors, 2) general health and physical characteristics, and 3) psychological aspects within and between the three proposed acute LBP severity definitions.

## Methods

For this study, we used baseline data from an ongoing cohort study of adults investigating the biopsychosocial factors related to transitioning from new onset acute LBP to chronic LBP. Study participants were recruited from communities in and around Durham, NC and Kannapolis, NC between February 2022 and November 2022. We utilized a community-based approach to recruit potential participants at both sites; this included advertisements on social media, newspaper articles, volunteer registries, emails distributed through university networks, flyers posted in or around the communities, and word of mouth. Recruitment in Kannapolis was primarily based on the MURDOCK Study. [5] The MURDOCK Study is a 12,526-participant community-based longitudinal cohort recruited from 2007-2013 (enrolled ∼2,000 participants per year) centralized in Cabarrus County. In Durham, we recruited participants using social media, flyers at local events, and the Duke Health Volunteer Registry. This registry can be queried for inclusion and exclusion criteria related to age, comorbidities (cancer/autoimmune conditions), and recent surgery/trauma. It contains over 7,200 volunteers accessible by email, and among those 55% are healthy, 56% identify as White, 35% identify as Black, and 5% identify as Hispanic. This study was reviewed and approved by the institutional review board at the Duke University School of Medicine.

Individuals interested in participating contacted research coordinators, who explained the study and initiated a telephone script for screening. To be eligible for study participation, individuals were required to be adults (≥18 years old) with acute LBP (i.e., LBP that started <4 weeks before screening and ≥30 days without LBP before the date of acute onset). We excluded individuals with a current or previous history of systemic inflammatory or autoimmune conditions (i.e., rheumatoid arthritis, spondyloarthropathies, irritable bowel syndrome, multiple sclerosis, Guillain-Barré Syndrome, human immunodeficiency virus, lupus), cancer (other than skin cancer), lumbar spine surgery, low back trauma (e.g., motor vehicle accident, falls), and congenital or acquired spinal defect (e.g., scoliosis). We also excluded individuals who were pregnant. Eligible individuals received and signed an informed consent form electronically via Research Electronic Data Capture (REDCap)[28; 29; 43] or in person. All participants were provided a copy of their signed consent form and scheduled for a baseline in-person visit no later than 6 weeks from the date of LBP onset. Before in-person data collection, participants completed study questionnaires to assess pain intensity, interference, and duration; depression, anxiety, and social measures via an online questionnaire using REDCap or, if the participant preferred, by phone. Study staff reviewed procedures with participants at the start of the in-person data collection visit. Participants were compensated for each in-person data collection visit and completion of electronic questionnaires.

### Defining Acute Low Back Pain Categories

We operationalized acute LBP severity categories using three different definitions, similar to those definitions recently proposed for high-and-low impact chronic LBP.[19] Two relied on LBP frequency combined with either: 1) pain impact frequency (impact-based) or 2) pain intensity (intensity-based), as well as one based on 3) pain interference frequency (interference-based). The impact-based acute LBP severity categorization was based on the definition used by the Population Research working group of the US National Pain Strategy to determine high-impact[55], adapting questions for an acute LBP population. Participants responded to the following questions about their LBP: 1) “Since the onset of your pain, how often have you had pain? Would you say Never, Some Days, Most Days, or Every Day?” and 2) “Since the onset of your low back pain, how often has pain limited your life or work activities? Would you say Never, Some Days, Most Days, or Every Day?” We categorized participants who answered “Most Days” or “Every Day” to both questions as having high-impact acute LBP. **Figure 1-A** illustrates how we used the same questions to categorize participants who did not fall into the high-impact acute LBP group into either medium- or low-impact categories.

**Figure 1.**
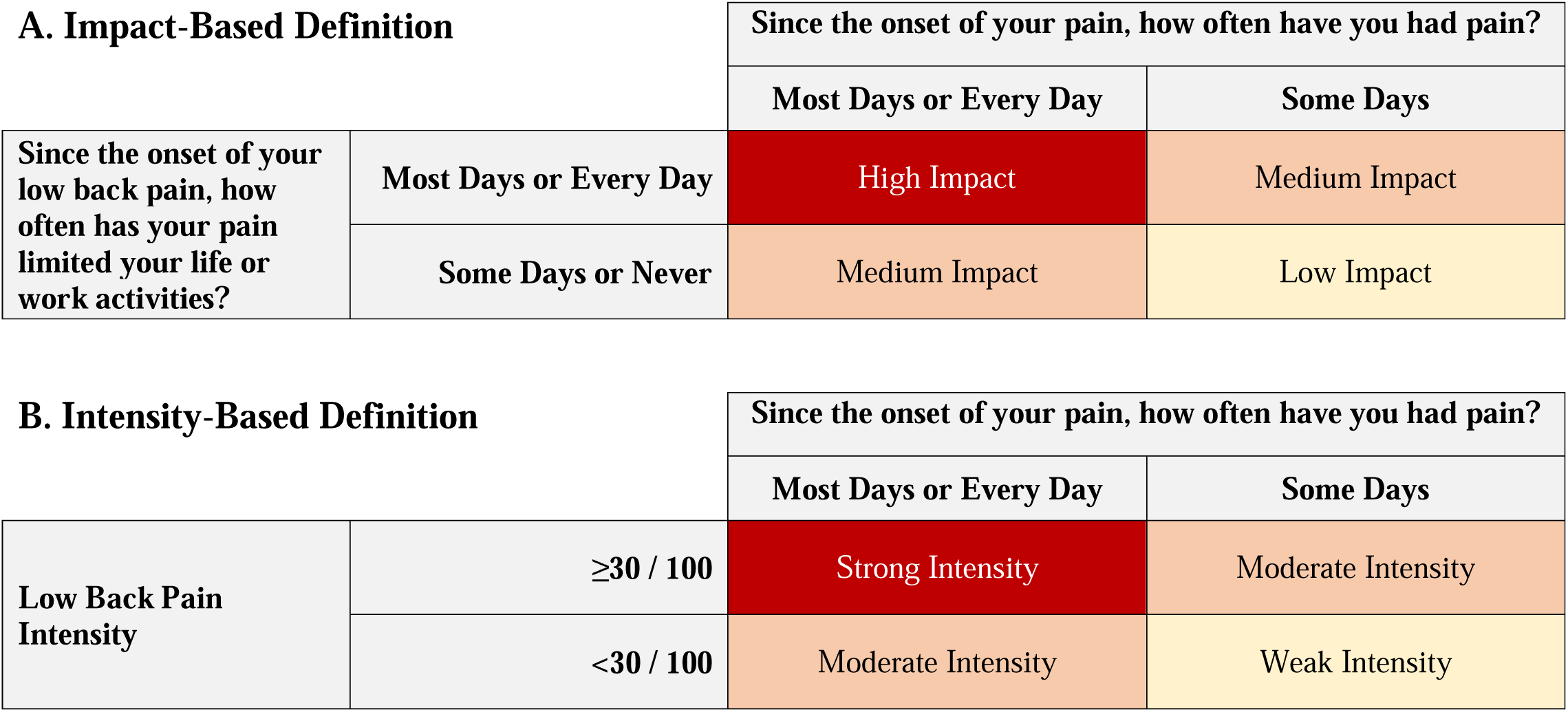
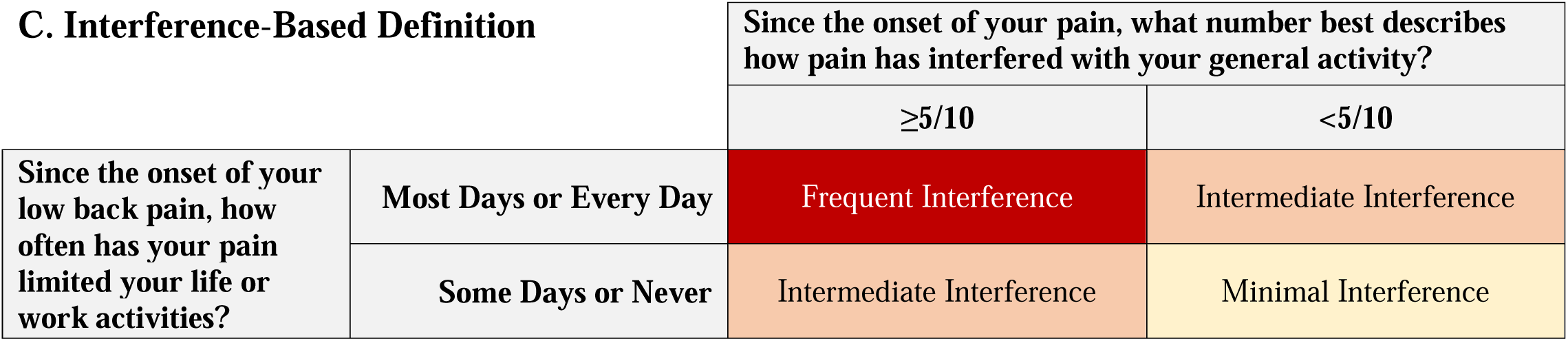
Acute Low Back Pain Severity Categorization Definitions.

Secondarily, the intensity-based severity definition was based on a combination of LBP frequency with LBP intensity using the 0-100 Visual Analogue Scale (VAS). Participants were categorized as having strong intensity acute LBP if they reported pain frequency as “Most Days” or “Every Day” and reported pain intensity ≥30 / 100. Two alternative cut-points were tested for pain intensity ≥20 / 100 and pain intensity ≥40 /100. These alternative intensity-based severity definitions and results are included in Supplementary Material A1. **Figure 1-B** illustrates how we categorized participants who did not fall into the strong-intensity acute LBP group into either moderate- or weak-intensity categories.

Finally, the interference-based severity definition included a measure of pain frequence and a measure of pain interference with the following two questions: 1) “Since the onset of your pain, how often have you had pain? Would you say Never, Some Days, Most Days, or Every Day?” and 2) the following PEG item: “Since the onset of your pain, what number best describes how pain has interfered with your general activity?” We categorized participants who for the first question answered “Most Days” or “Every Day” and reported an interference of ≥5/10 as having frequent interference acute LBP. **Figure 1-C** illustrates how we used the same questions to categorize participants who did not fall into the frequent-interference acute LBP group into either intermediate- or minimal-interference categories.

Here we present the results for the impact-based and the intensity based definition with the cut off value of 30/100 on the VAS. We elected to report these, as the alternative definitions had greater variability and did not perform as well.

Hedges g and Prevalence Ratio results for the alternative intensity-based definitions (VAS-20 and VAS-40) and the interference-only-based definitions are presented in Appendices B1-C6.

### Acute Low Back Pain History and Sensory Characteristics

Beyond the questions used to categorize acute LBP severity, we collected information regarding other pains and LBP treatments used since the onset of current LBP (adapted from the NIH recommended minimum dataset for chronic LBP),[16] personal history of prior LBP episodes, and family history of LBP and chronic pain. We also collected the STarT Back Screening Tool (SBT)[32], a 9-item questionnaire about LBP spreading down the leg(s), pain in other areas, pain interference with enjoyment of life (from the PEG),[40] pain interference with general activities (from the PEG),[40] perception of physical activity safety, pain catastrophizing, and LBP-related functional limitations to categorize individuals with LBP into low-, medium-, and high-risk categories based on their predicted risk of acute-to-chronic LBP transition.

We measured pain pressure threshold (PPT) at the upper trapezius (PPT-UT) and posterior superior iliac spine (PPT-PSIS) bilaterally using a standard rubber-tip algometer. All PPT tests were performed with participants seated with their hands in their lap. Study staff explained the procedure to the participants and instructed them to say “pain” as soon as the pressure applied first produced a painful sensation. Pressure was applied at a rate of 1-kgf/cm2/second until the participant’s PPT was reached up to 10.1 kgf. If the PPT was not reached before this ceiling, the measurement was recorded at a truncated value of 10.1 kgf. Three measurements for PPT-UT and PPT-PSIS were recorded on each side (alternating between left and right); we present the mean value (in kgf) of the PPT-UT and PPT-PSIS measurements.

### Sociodemographic Characteristics

We collected self-reported sociodemographic characteristics, including age, sex at birth, gender identity, racial identity, Hispanic ethnicity (yes/no), highest educational attainment, insurance type, marital status, and employment status. Participants could indicate one or more racial identities out of the following categories: American Indian or Alaskan Native, Asian, Black or African American, Native Hawaiian or Other Pacific Islander, or White. Participants also had the option to select “other” and provide a write-in racial category option or select “unknown” or “choose not to respond.” To assess racialized pain inequities in our impact-based, severity-based, and interference only-based categorization approaches, we grouped participants into two groups based on racialized sociopolitical positions: 1) “racially minoritized”, which included participants who identified as (regardless of ethnicity) Asian, Black or African American, and those who had selected more than one race category (there were no participants who identified as American Indian or Alaskan Native, or Native Hawaiian or Other Pacific Islander), and 2) “racially advantaged”, which included participants who identified as (regardless of ethnicity) White only.[6] Ethnicized pain inequities were examined based on whether participants indicated self-identifying as having Hispanic ethnicity or not. Participants reported sex at birth (female/male) and gender identity (cisgender, transgender, non-binary, genderqueer, agender, or gender fluid) using standardized items.[3]

### Social Health and Wellbeing

We captured two components of social health: social isolation and the ability to perform social roles. We ascertained social isolation using the Social Network Index (SNI)[4] , as recommended by the Institute of Medicine (IOM, currently knonwn as the National Academy of Medicine, [NAM]).[24; 44] The SNI asks questions about the frequency of contact with friends and family, religious participation, group membership, and marital status to classify individuals into four mutually exclusive groups: most isolated, very isolated, somewhat isolated, and not isolated.[24; 30] We used an item assessing the performance of social activities and roles collected from the Patient Reported Outcomes Measurement Information System (PROMIS) Scale v1.2 – Global Health to measure social function on a scale of 1-5 (where 1=Poor and 5=Excellent).[30]

### General Health and Clinical Characteristics

We used the General Health Item from the PROMIS Scale v1.2 – Global Health[30] to capture self-reported health status, classified into five mutually exclusive groups: Excellent, Very Good, Good, Fair, or Poor. In addition to general health, we asked participants about prior infections with the novel coronavirus disease of 2019 (COVID-19). Participants who indicated any known previous COVID-19 infections were also asked about the cumulative number of separate COVID-19 infections experienced and whether they considered their back pain to be related to a prior COVID-19 infection. Participant height and weight were collected in person at the study visit with a standard electronic scale and used to calculate body mass index (BMI).

### Health Behaviors

We classified participants into three mutually exclusive physical activity groups (0 min/week = inactive, 1-149 min/week = insufficiently active, and ≥150 min/week = sufficiently active) based on their responses to two questions: 1) “On average, how many days per week do you engage in moderate to strenuous exercise (like walking fast, running, jogging, dancing, swimming, biking, or other activities that cause a light or heavy sweat)?” and 2) “On average, how many minutes do you engage in exercise at this level?”[12; 24] We captured sleep disturbance using the PROMIS Short Form v1.0 – Sleep Disturbance 4a, where lower scores indicate better sleep (lowest possible raw score is 4; the highest possible raw score is 16).[57]

To categorize smoking status, we asked participants, “Have you smoked at least 100 cigarettes in your ENTIRE LIFE?” We classified individuals answering “no” to the first question as never smokers. Participants responding “yes” to the first question were asked a follow-up question: “Do you NOW smoke cigarettes every day, some days, or not at all?” Those responding “not at all” we classified as former smokers, while those responding otherwise were considered current smokers.[24] We ascertained substance use with two questions: 1) “Have you ever been drunk or used drugs more than you meant to?”; and 2) “Have you felt you wanted or needed to cut down on your drinking or drug use?” Response options for both questions were Never, Rarely, Sometimes, or Often.[16]

### Psychological Characteristics

To capture depressive symptoms, we collected the PROMIS Short Form v1.0 – Depression 4a.[46; 47] We collected stress symptoms using the IOM-recommended validated single-item question: “Stress means a situation in which a person feels tense, restless, nervous, or anxious, or is unable to sleep at night because their mind is troubled all the time. Do you feel this kind of stress these days?”[24] Participants are given the following response options: Not at All, A Little Bit, Somewhat, Quite a Bit, and Very Much.[20; 24] We used the Optimal Screening for Prediction of Referral and Outcome Yellow Flags (OSPRO-YF) 10-item questionnaire to estimate scores of other important psychological measures. The OSPRO-YF is a validated tool that provides predicted scores on the following measures: Fear-Avoidance Beliefs Questionnaire (both physical activity and work subscales; FABQ-PA and FABQ-W), Pain Anxiety Symptoms Scale (PASS-20), Pain Catastrophizing Scale (PCS), Patient Health Questionnaire-9 (PHQ-9), Pain Self-Efficacy Questionnaire (PSEQ), State-Trait Anxiety Inventory (STAI), State-Trait Anger Expression Inventory (STAXI), and Tampa Scale of Kinesiophobia (TSK-11).[9; 23; 41]

### Statistical Analysis

We separately summarized individual-level characteristics stratified by acute LBP severity categories for each categorization approach (i.e., impact-based, intensity-based, and interference only-based). Using low-impact acute LBP, weak-intensity acute LBP, and minimal-interference acute LBP categories as the reference groups, respectively, we present the relative magnitude of differences using prevalence ratios (PR) for categorical variables and Hedges’ *g* for continuous variables with their respective 95% confidence intervals (CI). Prevalence ratios were calculated as the proportion of the population in a given category, divided by the total reference population. Prevalence ratios were presented by comparing pairs of definitions to one another (i.e., impact-based vs. intensity-based, intensity-based vs. interference only-based, and interference only-based vs. impact-based). Hedges’ g effect sizes were presented for each severity definition individually. Interpretation of Hedges’ *g* is like Cohen’s *d* – with values around 0.2, 0.5, and 0.8 or more interpreted as small, moderate, and large differences between acute LBP groups.[31] We conducted all analyses in SAS 9.4 (Cary, NC) and used RStudio for data visualization.[36; 49; 56]

## Results

We screened 384 potential participants, 184 of whom met the study criteria. The most common reason for non-eligibility 158/200 (79%) was not meeting the study definition of acute LBP (i.e., duration of LBP >4 weeks at screening or reported LBP within 30 days before the current LBP episode), followed by history of cancer 16/200 (8%), active systemic inflammatory condition 11/200 (5.5%), history of lumbar surgery 7/200 (3.5%) and other reasons 8/200 (4%). Of those who screened eligible, 143/184 (77.7%) enrolled in the study, and 131/143 (91.6%) of those enrolled provided baseline data. No significant differences in the distribution of age, racial identity, or sex at birth were found between those eligible and did not enroll and those that did enroll in the study or those that did and did not provide baseline data. The most common employment status was working full-time (42.7%), with 42.0% of all participants reporting having some college, and 51.2% having a bachelor’s degree or higher educational attainment. We present sociodemographic characteristics for the overall sample in **Table 1**.

**Table 1.** Sociodemographic Characteristics of the Cohort.

|  | Total<br>(N=131) |
| --- | --- |
| **Age Category (years), n (%)** |  |
| 18-24 | 3 (2.3%) |
| 25-34 | 6 (4.6%) |
| 35-44 | 14 (10.7%) |

**Table 1.** Sociodemographic Characteristics of the Cohort
|  | Total<br>(N=131) |
| --- | --- |
| 45-54 | 30 (22.9%) |
| 55-64 | 36 (27.5%) |
| 65-74 | 29 (22.1%) |
| 75+ | 13 (9.9%) |
| **Female Sex At Birth**, n (%) | 77 (58.8%) |
| **Hispanic Ethnicity (yes)**, n (%) | 8 (6.1%) |
| **Racial Identity**, n (%) |  |
| Asian | 3 (2.3%) |
| Black or African American | 30 (22.9%) |
| White | 89 (67.9%) |
| Multiple Races | 6 (4.6%) |
| Unknown | 3 (2.3%) |
| **Employment Status**, n (%) |  |
| Working now, full-time | 56 (42.7%) |
| Working now, part-time | 18 (13.7%) |
| Looking for work, unemployed | 1 (0.8%) |
| Sick leave or maternity leave | 1 (0.8%) |
| Disabled for reasons other than back pain | 4 (3.1%) |
| Student | 3 (2.3%) |
| Retired | 40 (30.5%) |
| Keeping house | 3 (2.3%) |
| Other | 3 (2.3%) |
| Choose Not to Answer | 2 (1.5%) |
| **Highest Educational Attainment**, n (%) |  |
| High School Graduate, GED, or less | 9 (6.9%) |
| Limited College Education, Trade / Technical School Training, or Associates Degree | 55 (42.0%) |
| Bachelor's Degree or Some Graduate School but No Degree | 36 (27.5%) |
| Graduate Degree | 31 (23.7%) |
| **Marital Status**, n (%) |  |
| Single Never Married | 21 (16.0%) |
| Divorced | 19 (14.5%) |
| Separated | 4 (3.1%) |
| Widowed | 4 (3.1%) |
| Married / Living with Partner | 83 (63.4%) |

When considering the intersection between impact-based and intensity-based categorization approaches, 28.2% (95% CI: 20.5% to 36.0%) of participants were in both the low-impact and the weak-intensity categories, while 11.5% (95% CI: 6.0% to 16.9%) were in both the medium-impact and moderate-intensity categories and 5.3% (95% CI: 1.5% to 9.2%) were in both the high-impact and strong-intensity categories. The remaining 55% of the participants had different combinations of acute LBP categories across the two approaches, typically with a higher-tier intensity-based category than their impact-based category.

### Impact-Based Definition

The proportion of individuals in each impact-based acute LBP severity category decreased in a stepwise fashion from low-impact to high-impact acute LBP, with 52.7% (95% CI: 44.1% to 61.2%), 38.9% (95% CI: 30.6% to 47.3%), and 8.4% (95% CI: 3.6% to 13.1%) of our sample falling into low-, medium-, and high-impact acute LBP categories, respectively. We present results for differences in continuous variables in **Figure 2** comparing the medium- and high-impact acute LBP groups to the low-impact acute LBP group; PRs comparing the same groups to the low-impact acute LBP group are displayed in the next section along with PRs comparing strong- and moderate-intensity acute LBP groups to the weak-intensity acute LBP group in **Figure 4**. Compared to the low-impact acute LBP group, the medium-impact group was more likely to be female and less likely to report Hispanic ethnicity. When comparing the high-impact group to the low-impact acute LBP group, the high-impact acute LBP group was more likely to have leg pain related to their LBP. The low-impact acute LBP group had better PROMIS Global Physical Health scores than the medium- and high-impact groups. The medium- and the high-impact acute LBP groups were more likely to have medium risk and less likely to have low risk on the SBT. We could not calculate a PR to compare the likelihood of having high risk on the SBT because no participants in the reference group (low-impact acute LBP) had high risk on the SBT. Compared to those in the low-impact acute LBP group, those with medium- and those with high-impact acute LBP were also more likely to be in the racially minoritized group. Individuals in both the medium- and in the high-impact acute LBP groups were more likely to report associated leg pain and needing or wanting to cut back on alcohol or drug use than those with low-impact acute LBP. Interestingly, pain tolerance, as measured by pain pressure thresholds for the upper trapezius and posterior superior iliac spine, was higher for those in the low-impact group compared to those in the medium- and high-impact groups.

**Figure 2.**
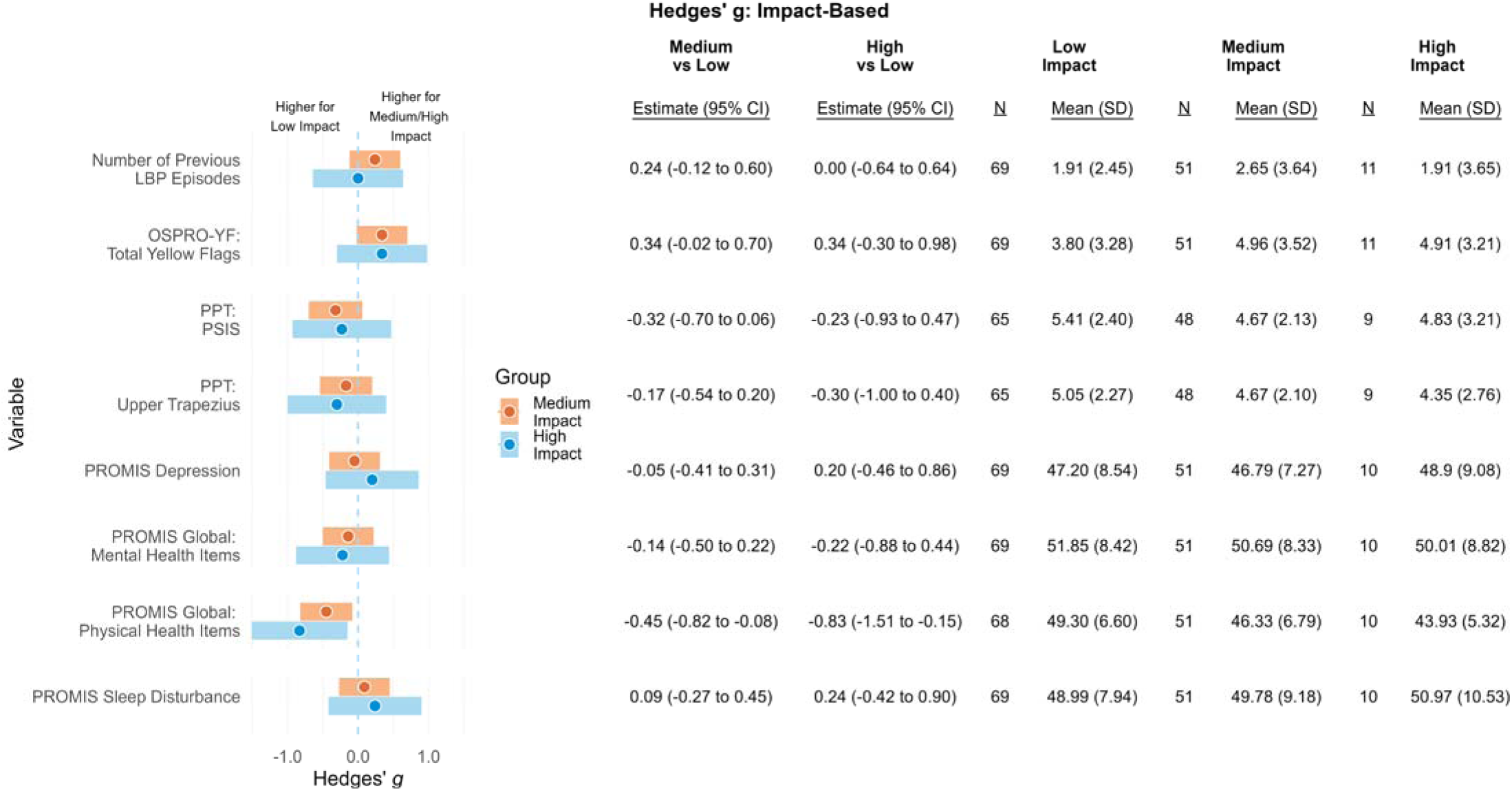
Hedges’ *g* for continuous variables comparing medium and high impacts to low impact acute LBP categories.

### Intensity-Based Definition

Compared to the impact-based acute LBP severity categorization, the proportion of our sample in each of our intensity-based acute LBP severity categories was more evenly distributed, with 33.6% (95% CI: 25.5% to 41.7%), 37.4% (95% CI: 29.1% to 45.7%), and 29.0% (95% CI: 21.2% to 36.8%) of our sample being in the strong-, moderate-, and weak-intensity acute LBP severity categories. For comparisons across intensity-based categories, we present the differences for continuous variables in **Figure 3**, comparing the moderate- and strong-intensity acute LBP groups to the weak-intensity acute LBP group; PRs comparing the same groups to the weak-intensity acute LBP group are in **Figure 4** (displayed alongside the medium- and high-impact acute LBP groups compared to the low-impact acute LBP group).

**Figure 3.**
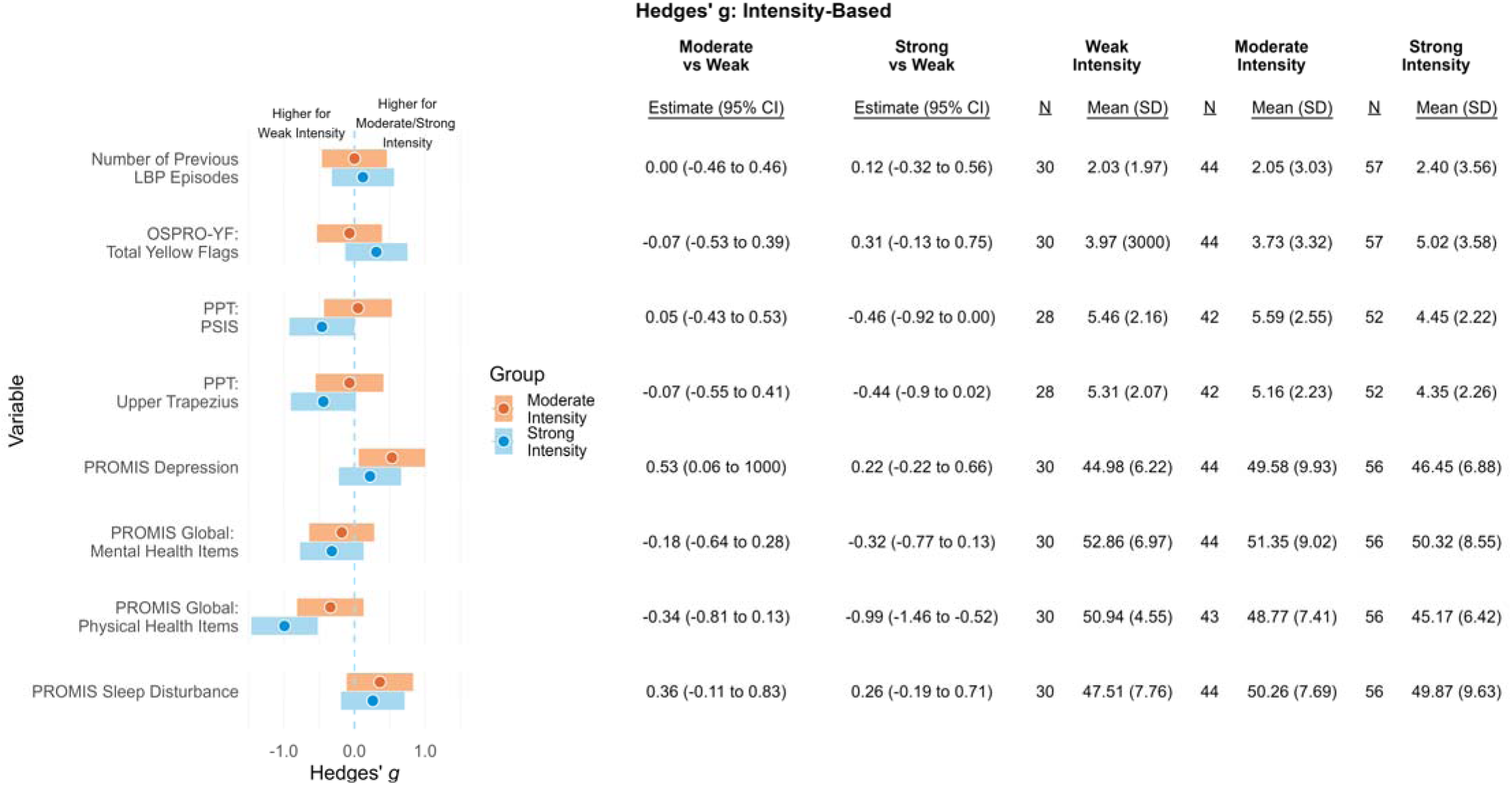
Hedges’ *g* for continuous variables comparing moderate-intensity to weak-intensity acute LBP categories.

**Figure 4.**
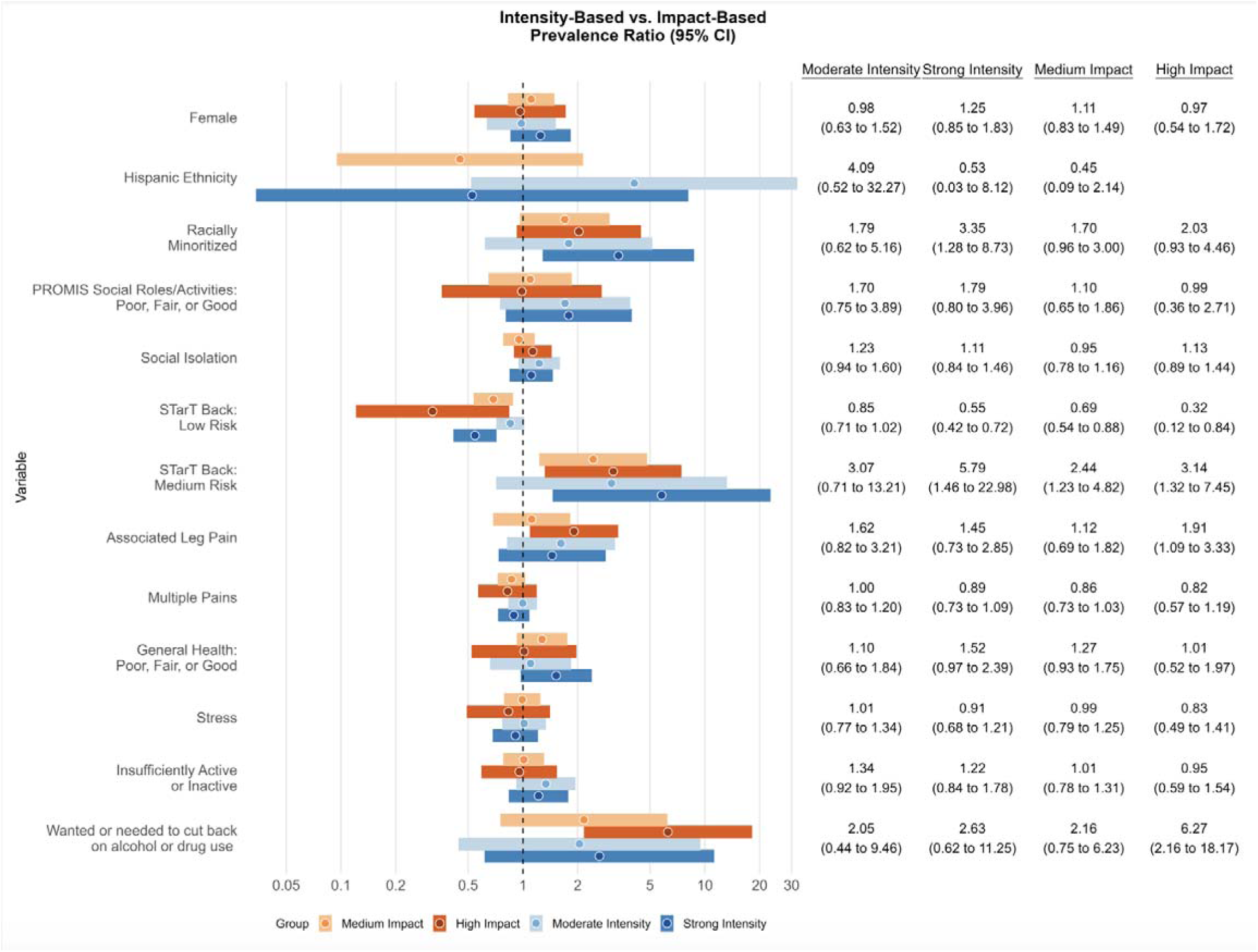
Prevalence ratios for categorical variables, comparing impact-based and intensity-based (with VAS-30/100 cut off) acute LBP severity categories. *Note each category is compared to its own definition’s referent category (i.e., moderate- vs. weak-intensity, strong- vs. weak-intensity, medium- vs. low-impact, and high- vs. low-impact).

Compared to the weak-intensity acute LBP group, the moderate-intensity acute LBP group was more likely to report social isolation, worse health, and worse ability to fulfill their usual social roles and activities. The strong-intensity group had lower PROMIS Global Physical Health scores relative to the weak-intensity acute LBP group and were more likely to have associated leg pain and worse general health. The group with strong-intensity acute LBP and the group with moderate-intensity acute LBP were more likely to have a medium risk of poor long-term outcomes on the SBT than the weak-intensity acute LBP group. In contrast, the strong-intensity acute LBP group was less likely to have a low risk on the SBT than the weak-intensity acute LBP group. Compared to the weak-intensity acute LBP group, the other two intensity-based groups were more likely to be in the racially minoritized group. The moderate- and strong-intensity acute LBP groups also reported a worse ability to complete their usual social roles and activities and reported wanting or needing to cut back on alcohol or drug use. Similar to the impact-based definition, pain tolerance was found to be higher for those in the weak-intensity group than for those in the moderate- and strong-intensity groups.

## Discussion

In this study, we have shifted from theoretical frameworks into empirical testing of different characterization approaches of acute LBP severity. We characterized acute LBP severity with participants from a community setting according to an impact-based definition, an intensity-based definition, and an interference only-based definition. The interference-only-based definition did not capture as much variability between acute LBP severity categories and usable information compared to the other two definitions. As such, the impact-based and intensity-based definitions provide a better understanding of domains that may contribute to or better predict the development of chronic LBP. The currently widely used characteristic for defining acute LBP is duration, with most studies indicating <6 weeks to represent the acute phase of LBP.[39; 53],[27] Some have used LBP intensity as an exclusion criterion, with individuals reporting intensities of <4/10 being considered ineligible.[35] These cut-offs are commonly used to exclude participants due to the potential inability to show important improvements in pain. This type of exclusion can lead to potential selection biases and an inability to characterize acute LBP accurately due to ceiling effects.[13; 35] Identifying new ways to systematically define acute LBP severity may help to identify specific sub-groups where treatments may be best tailored for response or to identify groups at high risk for the transition to chronic LBP. To our knowledge, such a characterization of acute LBP severity has never been published, and this study provides support for classifying acute LBP to capture differences among individuals with LBP. We recognize that our acute LBP impact-based definition was derived from a similar operational definition approach designed to measure the impact of activity-limiting chronic pain. While prior studies have proposed frameworks for which acute pain characterizations could be characterized[19], we are unaware of other studies that have examined the distribution of characteristics within proposed definitions for acute pain categorization. As such, one of our goals was to assess differences in sociodemographic, general and physical health, as well as psychological characteristics in acute LBP. Additionally, our intensity-based severity definition approach combined commonly used clinical trial eligibility criteria (≥4/10 pain intensity) and pain frequency to categorize acute LBP based on key characteristics.

Categorizing acute LBP severity using one or both of these proposed approaches may provide opportunities to assess their value in risk stratification and predicting the transition to chronic LBP. This stratification could help to inform interventions that improve care access and/or utilization for those who may be at the highest risk of chronic LBP but may not have accessed care.

Interestingly, there was little overlap in the across-category differences of participant characteristics between impact-based and intensity-based acute LBP severity categorization approaches, suggesting impact-based and intensity-based severity categorizations identified unique constructs associated with acute LBP. Differences were found between the two categorization approaches, primarily observed regarding their relationships to social constructs (i.e., sociodemographics, social roles, and social isolation). To our knowledge, reporting social constructs in the characterization of acute LBP severity is relatively rare. Social factors are highly complex and can be challenging to measure, which may contribute to the limited understanding of how social factors contribute to an individual’s experience with acute LBP. Two components of social health[7] (social isolation and the ability to perform social roles/activities) were more likely to be higher among participants in the stronger intensity-based categories. This points to differences in how various social constructs relate to individuals’ experiences with acute LBP and may help explain some differences between the two acute LBP severity categorization approaches. It may also be an indication of how LBP, even in the acute stage, is disruptive to social roles especially when examined through the intensity-based categories. Racialized and ethnicized inequities exist related to chronic LBP (e.g., inequities in pain severity, frequency, and pain-related disability).[2; 10; 11; 42] They are driven by intersecting systems and practices of racial and ethnic minoritization, discrimination, and socioeconomic/sociopolitical disadvantage that can negatively impact the biopsychosocial health and well-being of racially and ethnically minoritized groups over individual lifetimes and across generations.[1; 22; 26] Importantly, these same systems and practices of racial and ethnic inequity create and maintain socioeconomic/sociopolitical advantage that can protect the biopsychosocial health and well-being of racially and ethnically privileged non-Hispanic White groups over individual lifetimes and across generations.[6] In both impact-based and intensity-based categorizations, being racially minoritized was associated with being in the higher severity categories for acute LBP (i.e. medium- or high-impact and/or moderate- or strong-intensity). Conversely (and contrary to expectation), self-report of Hispanic ethnicity was associated with lower severity categorization for LBP on the intensity-based definition (i.e. moderate-intensity compared to strong-intensity) and could therefore be a protective factor. The same can be seen in the impact-based definition, no one identifying as Hispanic ethnicity was categorized in the highest group (i.e., high-impact), although the overall proportion of participants of Hispanic ethnicity was lower than the State estimate.[8] These findings underscore the importance of considering the impact that social factors, especially racialized and ethnicitized identities, have on both acute and chronic LBP outcomes.

These differences found here in our severity subgroup approaches present opportunities for standardization of acute LBP severity categorization based on specific populations and consistency for defining acute LBP severity for future pooling of results. For example, when investigating social factors such as social isolation, ethnicity, and possibly social networks or culture, it may be more appropriate to utilize the intensity-based severity definition since this categorization demonstrated differences between the various intensity-based severity category’s effect sizes. This demonstrates the need for special consideration of the representation of specific social factors when studying acute LBP. Understanding how social constructs relate to acute LBP severity and our acute LBP severity categorization approaches (intensity-based and impact-based) help provide a more complete characterization of adults experiencing acute LBP. It may help fill the gaps related to better predicting the transition to chronic LBP in future research.[37; 51; 52]

Although differences were noted in participant characteristics (i.e., ethnicity, social health factors, general health, etc.) between the two primary proposed categorizations, several important similarities were observed in the strength of effect sizes between categorization approaches. Current acute LBP intensity was consistently higher among participants in worse categories when using the intensity- or impact-based approach. Higher acute LBP intensity has previously been identified as a predictor of transitioning to chronic LBP [17]. PPT was consistently higher in participants in the weak-intensity or low-impact categories, indicating that these categorizations may adequately capture differences seen in pain tolerance across acute LBP severities. SBT scores that indicate a medium risk showed similar strong relationships with worse categories across categorization approaches. Previous research among individuals seeking care for LBP has indicated that the SBT provides clinically relevant stratification of individuals with a higher risk of persistent chronic symptoms [32; 33] , and a recent large observational study enrolling individuals seeking care for acute LBP found that the SBT accurately identified those who transitioned to chronic LBP.[51] Similar to these studies among care-seeking individuals, our findings indicate a similar psychological burden on participants with acute LBP within the community (regardless of care-seeking status) that may allow for chronic LBP risk stratification.

This study has several strengths, including comprehensive psychosocial measurements and a community-based cohort sample. However, there are limitations to this study. First, the primary purpose of this study was to collect preliminary data to assess the feasibility and acceptability of collecting a comprehensive set of measures from acute LBP participants within the community and follow these participants longitudinally. Our sample size was limited, impacting the precision of our Hedges’ *g* and PR estimates. This was especially true when comparing the high-impact acute LBP category to the low-impact category because of the relatively low number of participants in the high-impact category.

Second, we relied on self-reported physical activity for these analyses, which may misclassify physical activity measurement relative to accelerometry. We fielded the use of accelerometers to measure physical activity objectively, but this was limited to a subset of participants and occurred later in the project. Third, although we excluded participants who did not have 30 consecutive days without LBP before the onset of their current LBP episode to restrict our cohort from those with potential recurrent acute LBP, some participants reported other LBP episodes within the past year (which occurred and resolved ≥30 days before the current LBP episode) in the full data collection. Some participants may meet the consensus definition for recurrent LBP, depending on the context surrounding these previous LBP episodes.[50] A final limitation is the cross-sectional design, which does not allow us to fully investigate which of these categorization approaches is most helpful for the prediction of the transition to chronic LBP. Despite these limitations, our study comprehensively characterizes acute LBP severity and participants characteristics within the community, an understudied but potentially important area of LBP.

## Conclusion

This study used two primary severity approaches to categorize and describe community-based adults experiencing acute LBP. As expected, both impact-based and intensity-based severity categorization approaches, based on LBP intensity and LBP interference, had similar measures with consistent within-approach differences, including low- and medium-risk categories on the SBT. However, there were higher between-category differences in social isolation, social roles, and stress in the intensity-based definition compared to the impact-based definition. Results from this study support a recently proposed framework and suggest that intensity- and impact-based definitions may be acceptable to categorize acute LBP severity. Importantly these data allow for investigators to tailor their acute LBP severity subgroups to the needs of a given research question and provide the opportunity for standard definitions to be applied in future studies that are not solely reliant on temporal definitions of acute LBP.

## Data Availability

All data produced in the present study are available upon reasonable request to the authors

