## Supplementary Material for "Characterizing Acute Low Back Pain in a Community-Based Cohort"

APPENDIX

| **A) Intensity-Based Definition (VAS-20)** | | **Since the onset of your pain, how often have you had pain?** | |
| --- | --- | --- | --- |
|  |  | **Most Days or Every Day** | **Some Days** |
| **Low Back Pain Intensity** | **≥20 / 100** | Strong Intensity | Moderate Intensity |
|  | **<20 / 100** | Moderate Intensity | Weak Intensity |

| **B) Intensity-Based Definition (VAS-40)** | | **Since the onset of your pain, how often have you had pain?** | |
| --- | --- | --- | --- |
|  |  | **Most Days or Every Day** | **Some Days** |
| **Low Back Pain Intensity** | **≥40 / 100** | Strong Intensity | Moderate Intensity |
|  | **<40 / 100** | Moderate Intensity | Weak Intensity |

APPENDIX A1. Two alternative intensity-based definitions with a pain intensity cut-off of (A) 20/100 and (B) 40/100

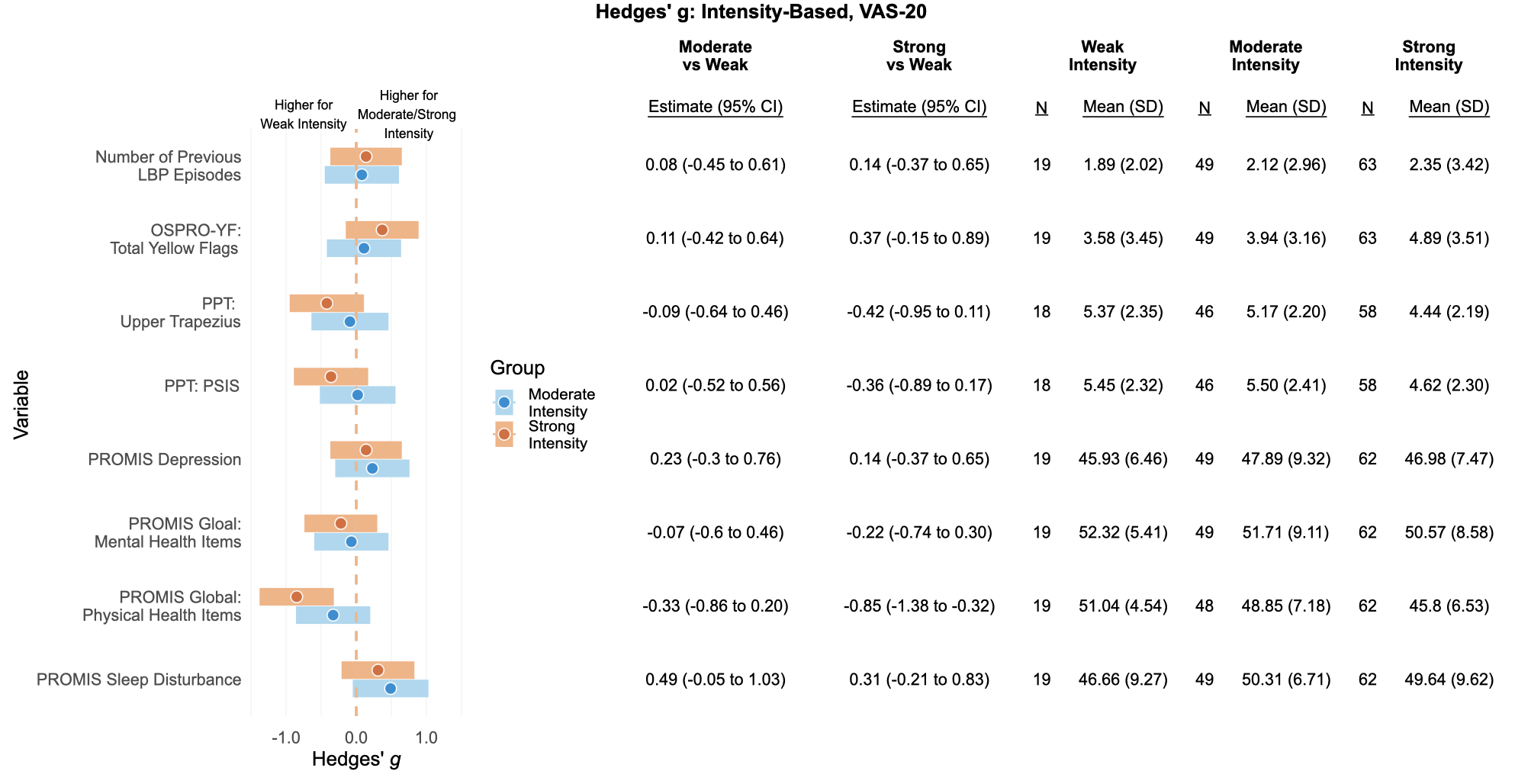

APPENDIX B1. Hedges’ *g* for continuous variables comparing moderate and strong intensity to weakinterference acute LBP categories in the Intensity-Based (VAS-20) definition.

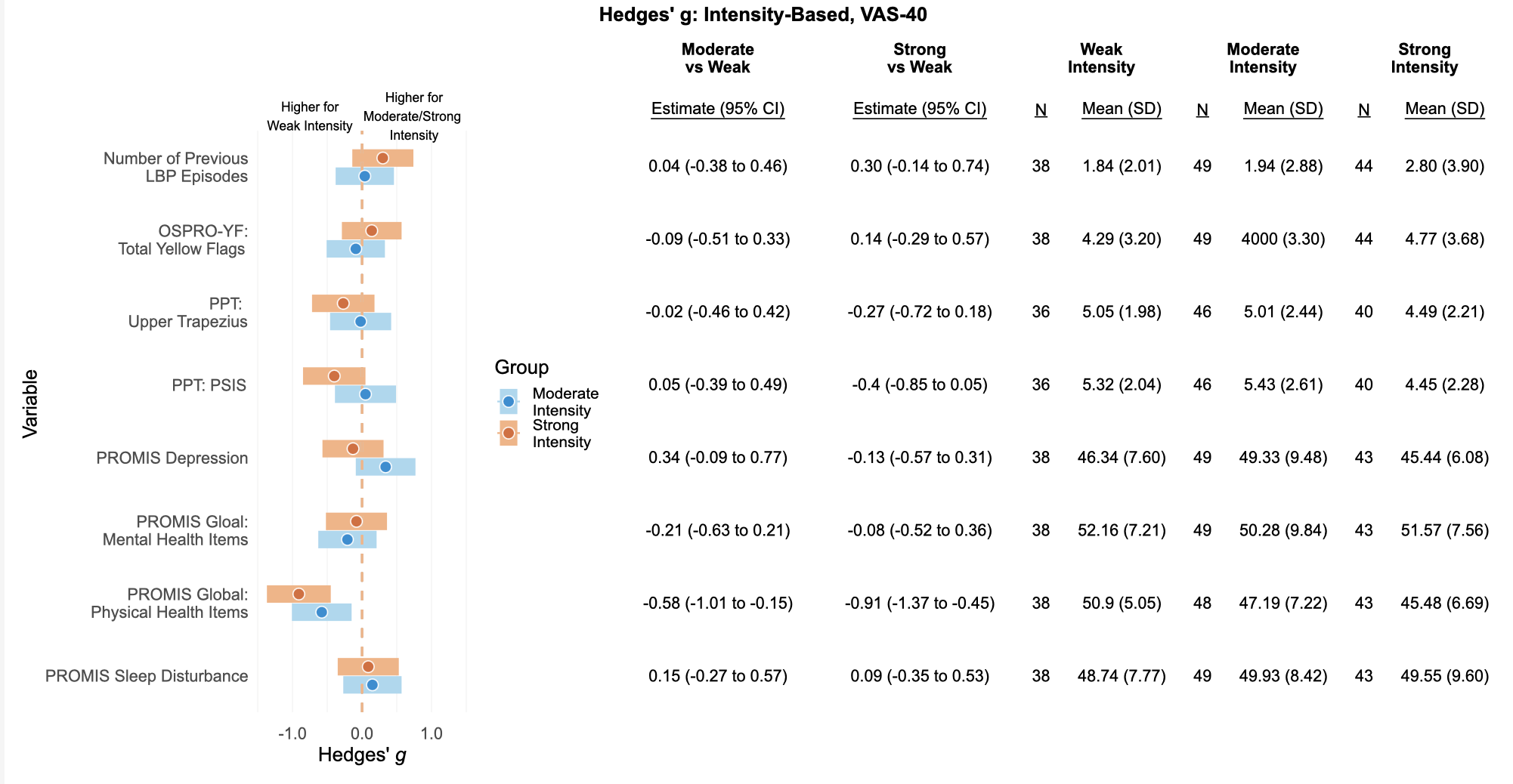
APPENDIX B2. Hedges’ *g* for continuous variables comparing moderate and strong intensity to weakinterference acute LBP categories in the Intensity-Based (VAS-40) definition.

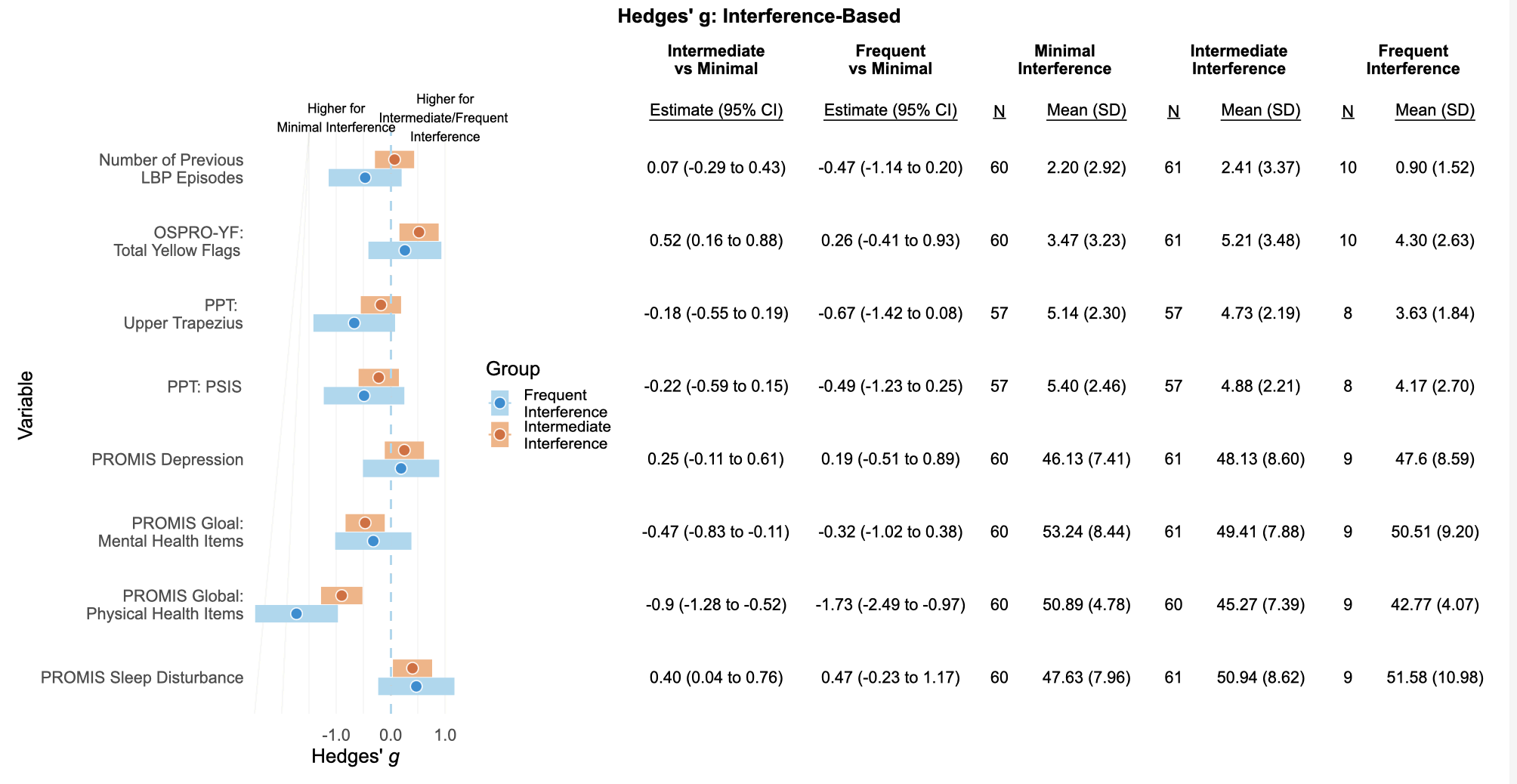

APPENDIX B3. Hedges’ *g* for continuous variables comparing intermediate and frequent intensity to minimal interference acute LBP categories.

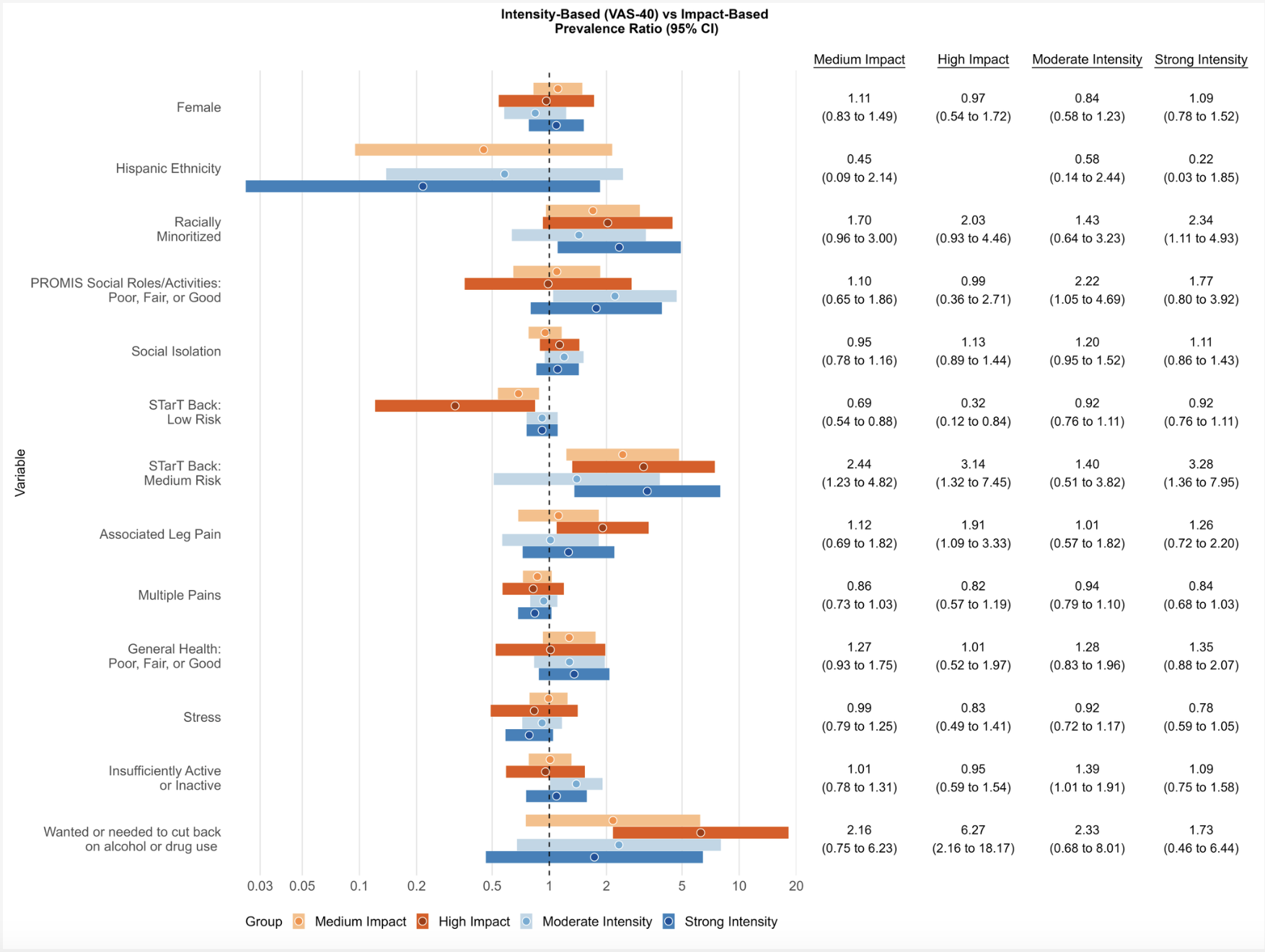
Appendix C1. Prevalence ratios for categorical variables, comparing impact-based acute LBP categories and intensity-based 40 acute LBP categories. *Note each category is compared to its own definition’s referent category (i.e., moderate vs. weak intensity, strong vs. weak intensity, medium vs. low impact, and high vs. low impact).

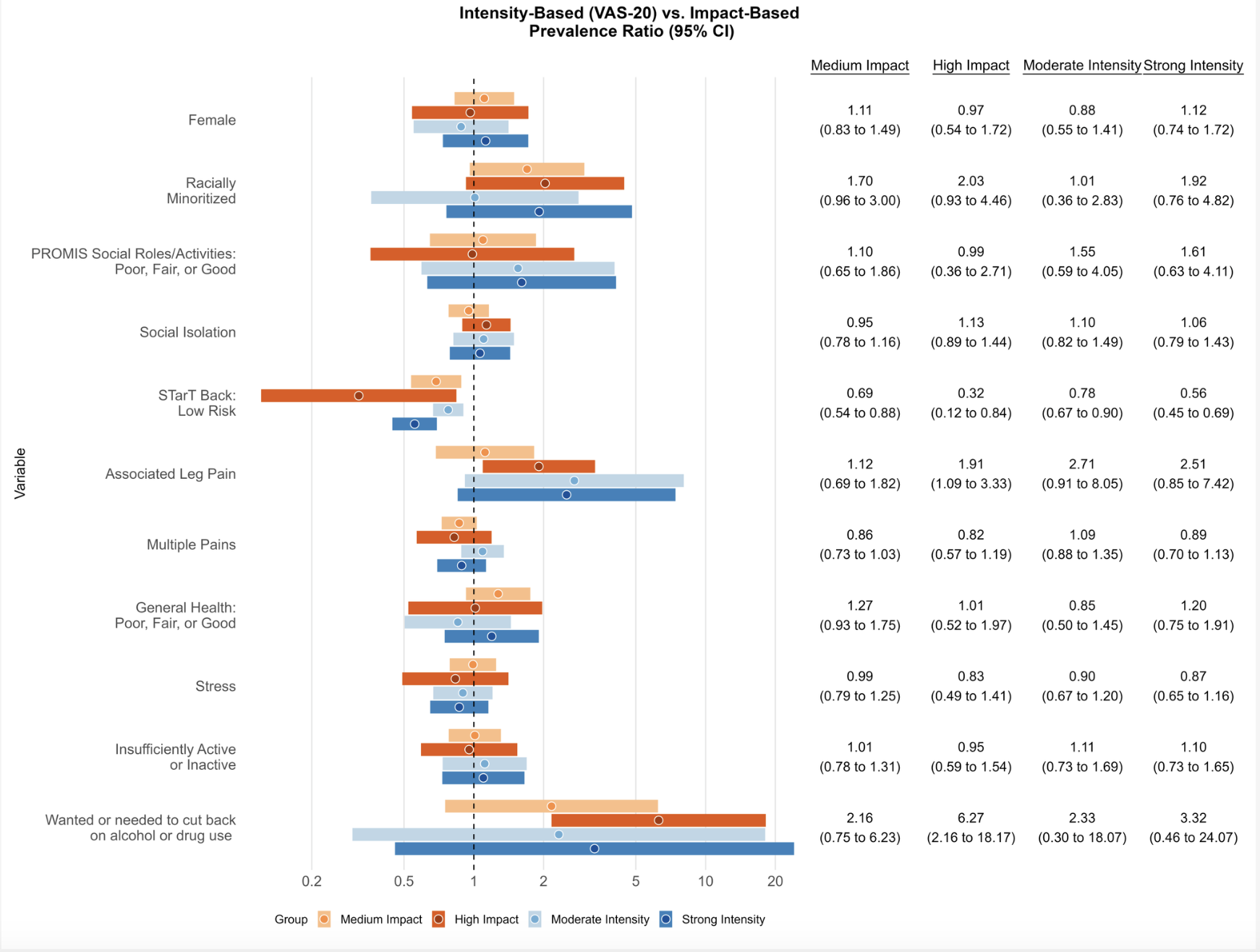
 Appendix C2. Prevalence ratios for categorical variables, comparing impact-based acute LBP categories and intensity-based VAS-20 acute LBP categories. *Note each category is compared to its own definition’s referent category (i.e., moderate vs. weak intensity, strong vs. weak intensity, medium vs. low impact, and high vs. low impact).

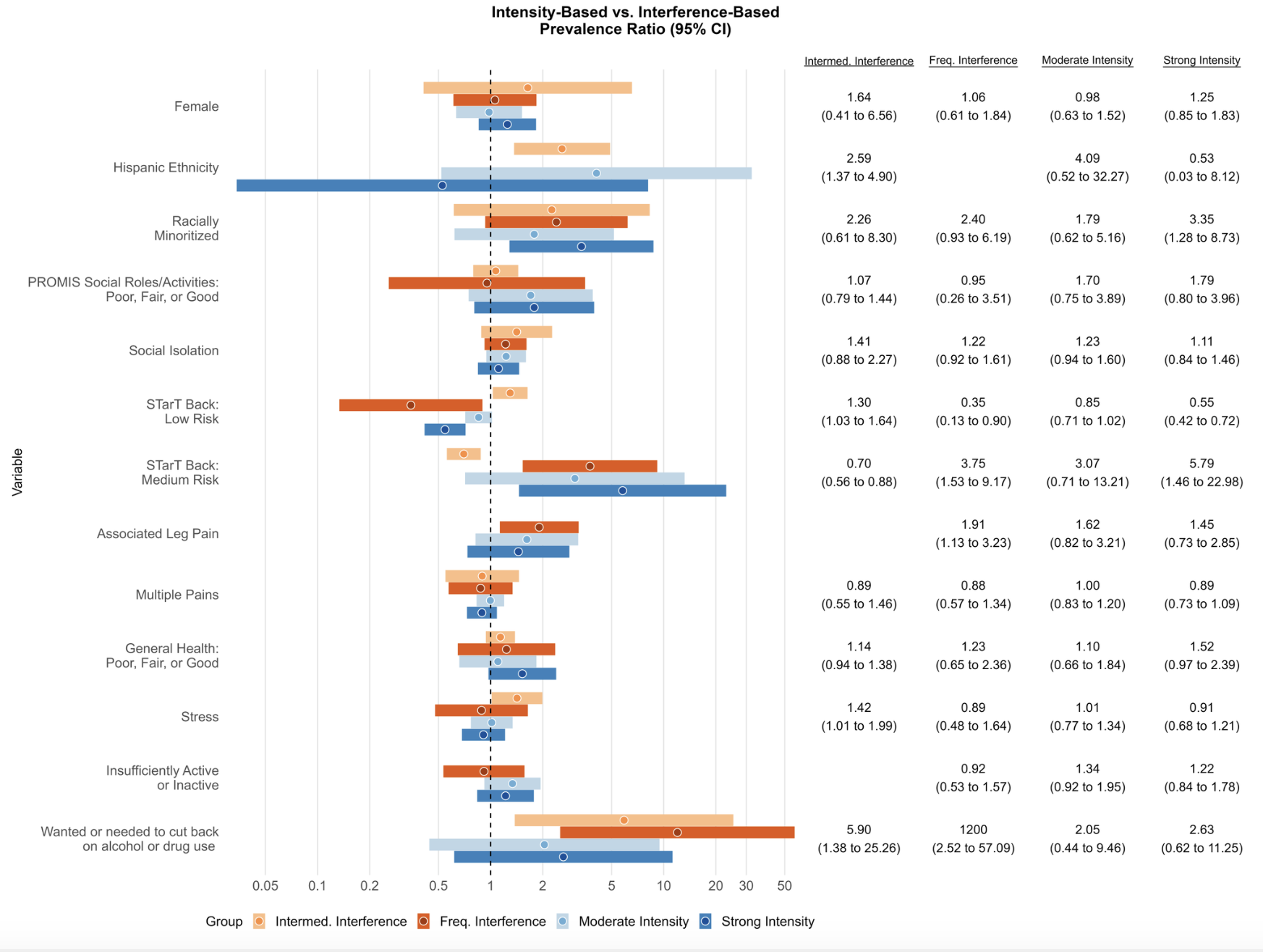

Appendix C3. Prevalence ratios for categorical variables, comparing interference-based acute LBP categories and intensity-based VAS- 30 acute LBP categories. *Note each category is compared to its own definition’s referent category (i.e., moderate vs. weak intensity, strong vs. weak intensity, intermediate vs. minimal interference, and frequent vs. minimal interference).

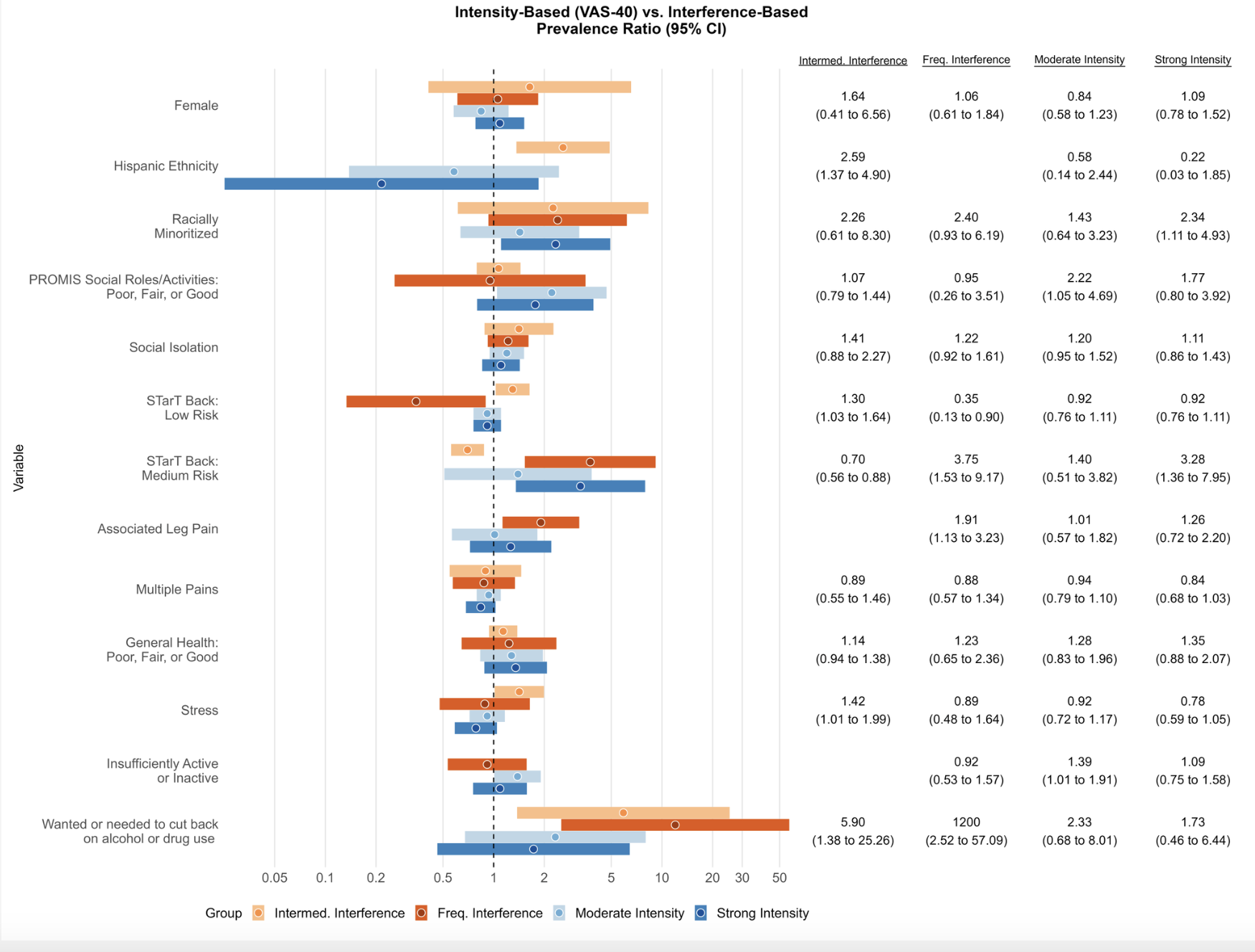

Appendix C4. Prevalence ratios for categorical variables, comparing interference-based acute LBP categories and intensity-based VAS-40 acute LBP categories. *Note each category is compared to its own definition’s referent category (i.e., moderate vs. weak intensity, strong vs. weak intensity, intermediate vs. minimal interference, and frequent vs. minimal interference).

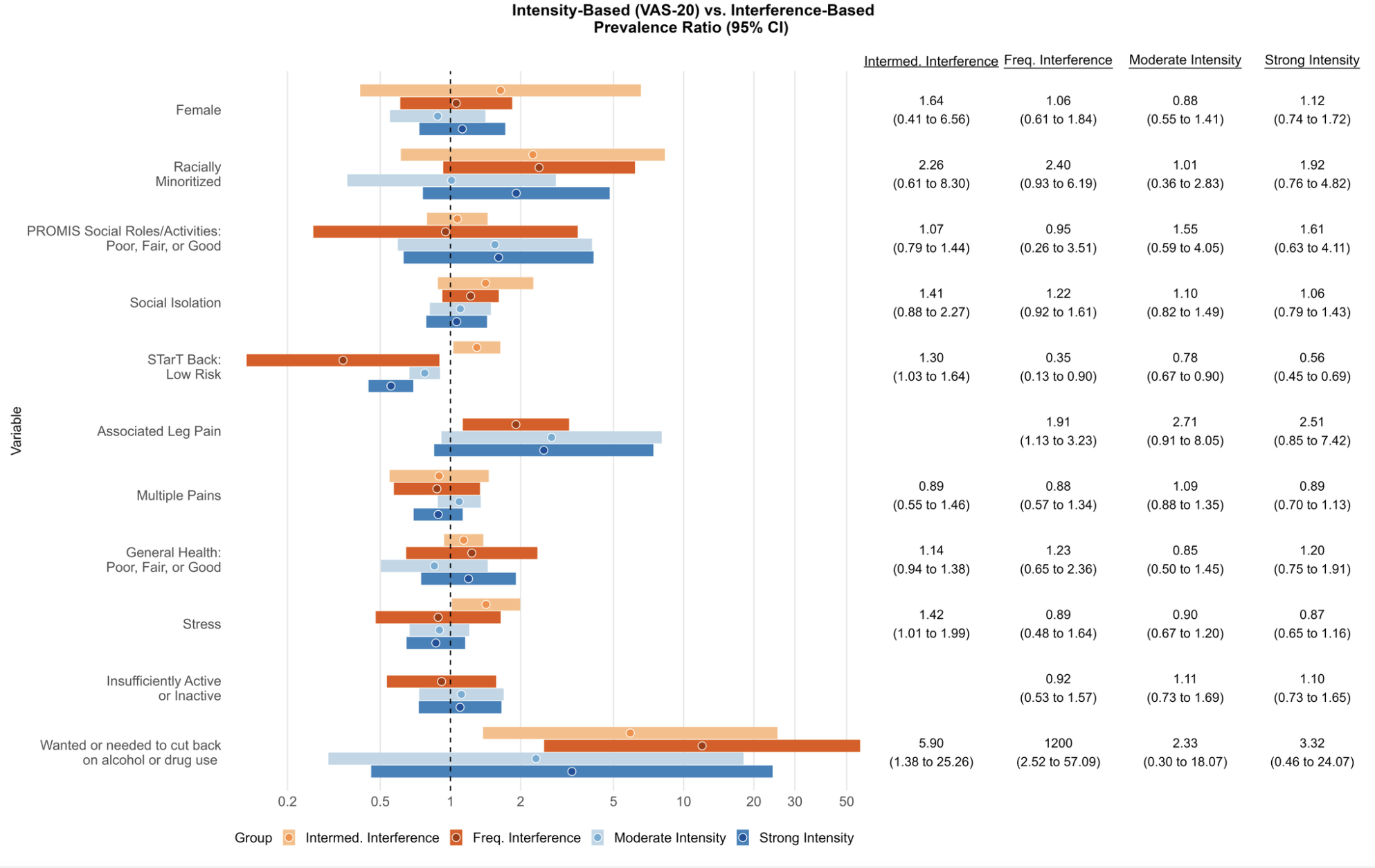

Appendix C5. Prevalence ratios for categorical variables, comparing interference-based acute LBP categories and intensity-based VAS-20 acute LBP categories. *Note each category is compared to its own definition’s referent category (i.e., moderate vs. weak intensity, strong vs. weak intensity, intermediate vs. minimal interference, and frequent vs. minimal interference).

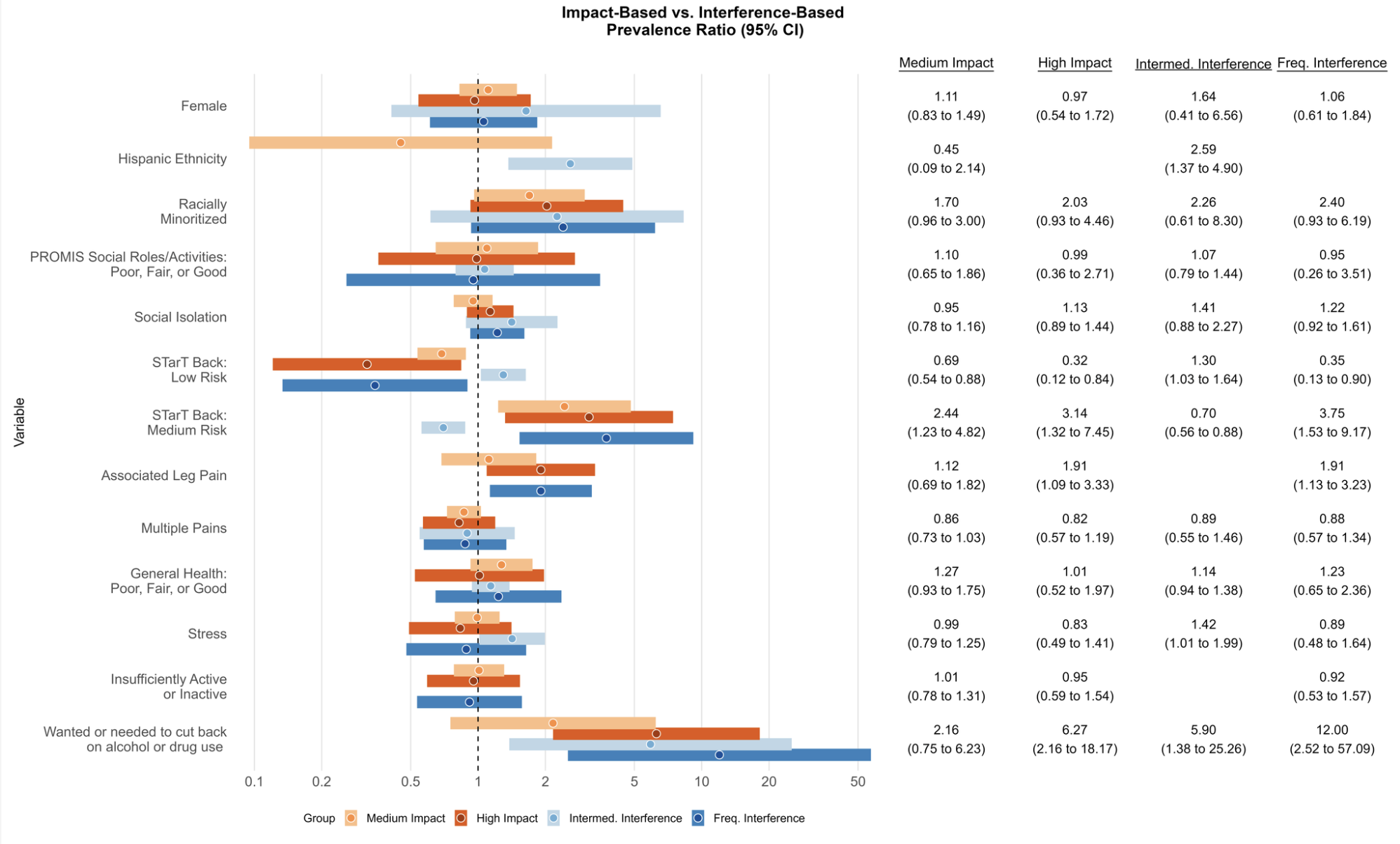
 Appendix C6. Prevalence ratios for categorical variables, comparing interference-based acute LBP categories and impact-based LBP categories. *Note each category is compared to its own definition’s referent category (i.e., , intermediate vs. minimal interference, and frequent vs. minimal interference, medium vs. low impact, and high vs. low impact).
